# The impact of brain lesion characteristics and the corticospinal tract wiring pattern on mirror movement characteristics in unilateral cerebral palsy

**DOI:** 10.1101/2020.01.31.20019893

**Authors:** Cristina Simon-Martinez, Ingar Zielinski, Brian Hoare, Lisa Decraene, Jacqueline Williams, Lisa Mailleux, Bert Steenbergen, Els Ortibus, Hilde Feys, Katrijn Klingels

**Affiliations:** KU Leuven - University of Leuven, Department of Rehabilitation Sciences, Leuven, Belgium; Institute of Information Systems, University of Applied Sciences Western Switzerland Valais (HES-SO Valais), Sierre, Switzerland; Behavioural Science Institute, Radboud University Nijmegen, Nijmegen, The Netherlands; Clinic for Pediatric and Adolescent Psychiatry, Psychosomatics, and Psychotherapy, Uniklinik RWTH Aachen, Germany; La Trobe University, School of Occupational Therapy, Victoria, Australia; Monash University, Department of Paediatrics, Victoria, Australia; College of Sport and Exercise Science, Institute for Health and Sport, Victoria University, Victoria, Australia; School of Psychology, Australian Catholic University, Melbourne, Australia; Centre for Disability and Development Research, Australian Catholic University, Melbourne, Australia; KU Leuven - University of Leuven, Department of Development and Regeneration, Leuven, Belgium; Rehabilitation Research Centre - REVAL, Faculty of Rehabilitation Sciences, Hasselt University, Diepenbeek, Belgium

**Keywords:** Cerebral Palsy, Upper Limb, Corticospinal Tract, Brain Damage, Mirror Movements

## Abstract

**Background:** Mirror movements (MM) influence bimanual performance in children with unilateral cerebral palsy (uCP). Whilst MM are related to brain lesion characteristics and the corticospinal tract (CST) wiring pattern, the combined impact of these neurological factors remains unknown.

**Objective:** To investigate the combined impact of neurological factors on MM.

**Methods:** Forty-nine children with uCP (mean age 10y6mo) performed a repetitive squeezing task to quantify similarity between MM activity (MM-similarity) and strength of the MM activity (MM-intensity). We used MRI to evaluate lesion type (periventricular white matter, N=30) cortico-subcortical, N=19), the extent of ipsilesional damage and damage to basal ganglia, thalamus and corpus callosum. The CST wiring pattern (17 CSTcontralateral, 16 CSTipsilateral, 16 CSTbilateral) was assessed with Transcranial Magnetic Stimulation. Data was analyzed with simple and multiple regression analyses.

**Results:** MM-similarity in the more-affected hand was higher with more damage to the corpus callosum. MM-intensity was higher in children with CSTcontralateral with damage to the basal ganglia and thalamus. In the less-affected hand, MM-similarity was explained by the interaction between lesion type and CST wiring pattern, with higher MM-similarity in children with cortico-subcortical lesions in the CSTcontralateral group. MM-intensity was higher with larger damage to the corpus callosum and unilateral lesions.

**Conclusions:** A complex combination of neurological factors influences MM characteristics and the mechanisms differ between hands.

## Introduction

One of the impairments that may hamper the performance of daily life activities in children with unilateral cerebral palsy (uCP) is the presence of mirror movements (MM)^1^. MM are described as involuntary movements in one limb, which are initiated by simultaneous, voluntary activation of the homologous muscles in the other limb^2,3^. MM are a physiological feature in typically developing children, gradually disappearing during the first decade of life^2^ due to the gradual maturation of the corpus callosum^4^.

In many children with uCP, MM are more pronounced and persistent^3,5,6^ and negatively impact the performance of asymmetric bimanual tasks and the time required to complete them^1^. Two hypotheses have been put forward as potential mechanisms of MM occurrence in uCP: (1) an ipsilateral corticospinal tract (CST) projecting from the contralesional motor cortex to both hands; and (2) insufficient interhemispheric inhibition between the two primary motor cortices^7^. The reorganization of the CST in three wiring patterns (i.e. contralateral (CSTcontra), ipsilateral (CSTipsi) and bilateral (CSTbilat), is unique to children with uCP due to the underlying brain lesion and refers to the efferent motor input to the affected hand^8,9^. The presence of an ipsilateral tract (either in CSTipsi or CSTbilat wiring pattern) has been found to result in more severe MM, although those with CSTcontra may also present MM^10–13^. To the best of our knowledge, there are no studies investigating the impact of interhemhispheric inhibition on MM in children with uCP. In children with bilateral CP, a simultaneous bilateral activation of both primary motor cortices and insufficient interhemispheric inhibition, together with decreased fiber integrity in the transcallosal pathways has been related to MM^14^. However, this mechanism may vary between types of CP and between hands.

Brain lesion characteristics such as the lesion type, extent and location and the presence of bilateral lesions could provide additional information to the underlying mechanism of MM occurrence. The *type of lesion*, related to the timing of the lesion during gestation, can be classified into three categories: malformations (1^st^ and 2^nd^ trimesters of pregnancy), periventricular white matter lesion (PV, early 3^rd^ trimester), and cortico-subcortical lesions (CSC, late 3^rd^ trimester and around birth)^15^. Klingels et al (2016) demonstrated that malformations and PV lesions were related to more severe MM in the more-affected hand than other lesion types^6^. However, Riddell et al (2019) found no difference between lesion groups in the more-affected hand^12^. These contrasting results highlight the need for further investigation, including the influence of other brain lesion characteristics.

Additional studies have reported a relationship between *lesion extent* and MM occurrence. The extent of the lesion in children with PV lesions has been related to MM occurrence, whereby a larger lesion was related to stronger MM^11^. Whether this relationship also applies to other lesion types i.e. malformations and CSC, remains unknown. Other brain lesion characteristics influencing MM, such as *lesion location* and the presence of *bilateral lesions*, have not previously been investigated.

Overall, a few studies suggest that neurological factors partially explain the MM phenomenon in children with uCP, although other factors may also play a role. In previous studies, the use of an observation-based scale producing ordinal data (i.e. Woods and Teuber^5^) hampers objective quantification of the MM activity. In this study, quantitative methods with advanced analyses that better identify different MM characteristics^16^ will provide deeper insights in the MM phenomenon. The aim of the study is to investigate the role of single neurological factors (i.e. brain lesion characteristics and CST wiring pattern), as well as their combined impact on MM characteristics in children with uCP, using a novel, objective method to quantify MM^16,17^. We hypothesize that the identification of the combined impact of the neurological factors will better explain MM occurrence in children with uCP.

## Methods

### Participants

Children with congenital uCP, aged between 5-15y were recruited from the CP care program of the University Hospital Leuven (Belgium) and Monash Children’s Hospital (Melbourne, Australia) from June 2015 until November 2017. Children with congenital uCP were included if they were sufficiently mentally capable and cooperative to understand and complete the test procedures. Children were excluded if they (1) received upper limb Botulinum toxin injections six months prior to testing, (2) underwent UL surgery two years prior to testing or (3) had MRI- or TMS-related exclusion criteria (i.e. shunt or any sort of metal in the body, not controlled epilepsy). All children verbally assented to participate in the study and all parents signed a written consent form prior to participation in accordance with the Declaration of Helsinki. The study was approved by the local Ethics Committees (Leuven: S55555 and S56513; Monash Health HREC: 12167B).

### Assessments

Children recruited at the University Hospital Leuven (Belgium) underwent Magnetic Resonance Imaging (MRI) for the purposes of this study. At Monash Children’s Hospital (Australia), the child’s most recent clinical MRI scan was used for this study. All participants underwent Transcranial Magnetic Stimulation (TMS) and MM testing using a quantitative device.

**Mirror movements** were quantitatively assessed using the same procedure for both sites following a previously published protocol^16^. The device consists of two grip-force transducers connected to a windmill. Children held the transducers between thumb, index and middle finger, although they could use more fingers if they had difficulties applying this pinch-grip with their more-affected hand (the grip of the less-affected hand was always matched to the grip of the more-affected hand). First, we measured the maximum voluntary contraction (MVC) of each hand three times and the average of the three repetitions was used for analysis. The less-affected hand was always measured first. Second, the children performed the task by repetitively squeezing one of the transducers with one hand, while the other hand just held the other transducer. The transducer that was squeezed actively was connected to a miniature windmill. Once the transducer was pressed beyond a threshold of at least 15% of the MVC and if the grip-force exceeded 1.5 kg, the windmill started rotating. By executing a repetitive squeezing pattern with a frequency of ≥ 1Hz, the child could keep the windmill turning. A trial consisted of a squeezing period of 5 seconds. The force profiles (mV/V) during 20 trails (10/hand) were collected using PsychoPy standalone version 1.82^17^ and further used to extract the MM characteristics (see Data analysis).

**Magnetic Resonance Imaging** was collected in Leuven (Belgium) with a 3T Philips Ingenia scanner (Philips Healthcare, Best, the Netherlands) with a protocol detailed elsewhere^18^. Briefly, we acquired a T1-weighted 3D fluid attenuated inversion recovery (3D FLAIR) image and a magnetization prepared rapid gradient echo (MPRAGE). Prior to data acquisition, children were familiarized with the scanner by performing scan-related tasks (e.g. laying still, wearing a helmet and earplugs). At Monash Children’s Hospital (Australia), existing 3DFLAIR and/or MPRAGE MRI images were retrieved from the medical records if they were collected after the age of 3 years, as the myelination process is then complete^19^.

Brain lesion characteristics (lesion location and extent) were identified by an experienced child neurologist (EO). Brain lesion type was classified into three groups: brain malformations, PV and CSC lesions^20^. The quantification of the brain lesion characteristics was performed with a valid and reliable semi-quantitative scale^21,22^ containing a graphical template of the lobes, corpus callosum cerebellum and subcortical structures. First, the MRI slices that corresponded to the template were identified and the lesion was drawn by a child neurologist experienced in the use of the scale (EO). We calculated the score of the total ipsilesional damage (total 0-17) based on the damage to the lobes (0–3 for each lobe, i.e., total of 0–12) and damage to the subcortical structures (0–5 including the lenticular, caudate, posterior limb of the internal capsule (PLIC), thalamus and brainstem). Lesion location was indicated by the damage to the frontal and parietal lobes (0–4), the basal ganglia and thalamus (0–3) and the corpus callosum (0-3, according to the number of thirds that were damaged). The presence of bilateral lesions or purely unilateral lesions was also documented.

#### Transcranial Magnetic Stimulation

TMS was used to identify the underlying CST wiring pattern with a previously described protocol^18^. We used a Magstim 200 Stimulator (The Magstim Company Ltd, Whitland, Wales, UK) with a focal 2×70mm figure-eight coil to evoke motor evoked potentials (MEPs) on the adductor pollicis brevis muscle in both hands. The child sat on a chair with both hands relaxed on a cushion. A cap was placed on the head on which a raster was drawn to help with the identification of the optimal location for evoking an MEP (≥ 50µV amplitude) for each hand (i.e. hotspot). We started with an intensity of 30% which was gradually increased until a MEP was evoked (resting motor threshold, RMT). Next, 10 MEPs were collected at 120% of the RMT. If the RMT was 85% or higher, 10 MEPs were collected at maximum stimulator output (100%). We first identified the CST from the non-lesioned hemisphere to the less-affected hand, followed by the ipsilateral CST (innervating the more-affected hand). Lastly, we stimulated the lesioned hemisphere to search for a remaining contralateral CST. This protocol indicated the CST wiring pattern, i.e., contralateral (CSTcontra, the affected hand receives input from the crossed CST, originating in the lesioned hemisphere), ipsilateral (CSTipsi, the affected hand receives input from the uncrossed CST, originating in the non-lesioned hemisphere), or bilateral (CSTbilat, the affected hand receives input from both the crossed and uncrossed CSTs, originating in the lesioned and non-lesioned hemispheres, respectively). There were no adverse events.

### Data analysis

#### Mirror movements

A custom-written script was used to analyse the MM data with the Psychopy standalone version 1.82 software. First, we visually inspected the 20 trials per child (10 trials with each hand). One trial displayed graphs of the grip forces of both hands. When such forces appeared in the passive hand, they were an indication of the presence of MM. Visual inspection of the trials was needed to identify the valid trials. A valid trial consisted of at least 10 squeezes of the active hand above a threshold of 1.5 kg and with a grip-force of 15% of the child’s MVC. The first 500ms after the ‘start’ signal were not considered to account for a delay in reaction time. Only valid trials were retained for the statistical analysis.

To quantify the MM, both the similarity (i.e. coupling of force profiles) and intensity (i.e. strength of the mirroring activity) were calculated, following the procedures described elsewhere ^16^. MM-coupling consisted of the maximum correlation-coefficient representing an index of similarity between the squeezing signals of both hands (range from 0-1, ‘0’ indicating no MM and ‘1’ mirroring activity identical to the voluntary movements), which was averaged across trials. Second, we calculated the MM-intensity by computing the mean grip force as the difference between the heights and troughs of the force signal and this was averaged across trials. The ratio between the passive and active hand was computed as follows: 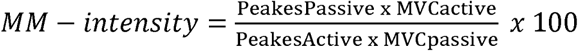. A higher MM-intensity ratio indicates more severe MM.

### Statistical analyses

First, descriptive statistics were used to document the distribution of MM characteristics according to the brain lesion characteristics and the CST wiring pattern. Second, we documented differences in MM data according to affected side, gender and its relation with age, using parametric statistics. Third, we investigated the association between *each* brain lesion characteristic and CST wiring pattern on MM characteristics in each hand using simple linear regression. Next, we explored to what extent MM characteristics in each hand could be explained by the brain lesion characteristics, and their interaction with the CST wiring pattern in a multiple regression analysis. This analysis was fitted with the backward elimination method until we identified a set of variables significantly contributing to the model, and R^2^ and p values are reported. Those factors that did not influence (p<0.05) any MM characteristic in any hand in the simple linear regression analyses were not included in the multiple linear regression model.

Statistical significance was set at α<0.05. All statistical analyses were computed with SPSS Statistics for Windows version 25.0 (IBM Corp., Armonk, NY).

## Results

### Participants

Eighty-six children were recruited for this study (mean age 10y 6mo, SD 2y 6mo, 43 girls, 45 right hemiplegia). We only included participants with complete datasets (MM data, brain lesion characteristics and CST wiring pattern), and we excluded malformations due to low incidence^23^, resulting in a sample of 49 children (mean age 10y 6mo, SD 2y 8mo, 25 girls, 24 right unilateral CP, Manual Ability Classification System (MACS): 18 MACS I, 18 MACS II and 13 MACS III). Reasons for exclusion were either related to the MM evaluation (n=2), TMS assessment (n=20), MRI session (n=11) or mixed reasons (n=4) and are described in detail in Supplementary Materials Table S1. Thirty children had a PV and 19 a CSC lesion; 30 had purely unilateral lesion and 19 a bilateral lesion. The CST wiring pattern was CSTcontra in 17 children, CSTipsi in 16 and CSTbilat in 16. Demographic and MM data did not differ between sites (Leuven, Belgium N=44, Melbourne, Australia N=5; Supplementary Materials Table S2).

MM characteristics did not differ according to affected side (MM-similarity, p>0.55; MM-intensity, p>0.52), gender (MM-similarity, p>0.23; MM-intensity, p>0.24) nor were related to age (MM-similarity, r=0.07 to 0.13, p>0.38; MM-intensity, r= −0.06 to 0.07, p>0.64).

### Individual influence of each neurological factor on mirror movements

Table 1 reports the MM data in each hand according to the CST wiring pattern and the brain lesion characteristics, as well as the association of each variable on the MM characteristics. Each factor related differently to MM-similarity and MM-intensity (Table 1). In both hands, the CST wiring pattern, damage to the corpus callosum, basal ganglia and thalamus and purely unilateral lesions were the factors that influenced most MM characteristics (p<0.05) with low to medium R^2^ (R^2^=0.08-0.25). Children with CST ipsi and purely unilateral lesions had higher MM-similarity and MM-intensity in both hands. Similarly, MM-similarity in both hands increased with more damage to the corpus callosum, with a similar pattern for MM-intensity in the less-affected hand. MM-intensity in both hands was stronger with damage to the basal ganglia and thalamus and followed a similar trend for MM-similarity in the more-affected hand. Damage to the frontal and parietal lobe and lesion extent did not influence the MM characteristics in any hand (p>0.05, R^2^=0-0.07), thus they were not included in the multiple regression analyses.

**Table 1.** Descriptive statistics (X (SD)) and simple regression analyses of MM similarity and MM intensity according to the CST wiring and the brain lesion characteristics.

|  |  | More affected hand |  |  |  | Less affected hand |  |  |  |
| --- | --- | --- | --- | --- | --- | --- | --- | --- | --- |
|  |  | MM-similarity |  | MM-intensity |  | Less affected hand SIM |  | Less affected hand INT |  |
|  |  | X (SD) | R<sup>2</sup> (p-value) | X (SD) | R<sup>2</sup> (p-value) | X (SD) | R<sup>2</sup> (p-value) | X (SD) | R<sup>2</sup> (p-value) |
| **CST wiring pattern** |  |  | **0.13 (0.04)\*** |  | **0.22 (0.003)\*** |  | **0.25 (0.001)\*** |  | **0.10 (0.09)** |
| Contralateral |  | 0.18 (0.19) |  | 24.85 (36.63) |  | 0.16 (0.19) |  | 18.04 (29.68) |  |
| Bilateral |  | 0.32 (0.26) |  | 38.16 (22.99) |  | 0.36 (0.25) |  | 45.69 (57.55) |  |
| Ipsilateral |  | 0.40 (0.29) |  | 66.59 (39.58) |  | 0.49 (0.29) |  | 52.67 (49.16) |  |
| **Lesion type** |  |  | **0.02 (0.40)** |  | **0.01 (0.41)** |  | **0 (0.91)** |  | **0.08 (0.047)\*** |
| Periventricular lesion |  | 0.27 (0.26) |  | 39.29 (39.51) |  | 0.34 (0.31) |  | 27.55 (37.32) |  |
| Cortico-subcortical lesion |  | 0.33 (0.26) |  | 48.42 (34.59) |  | 0.33 (0.23) |  | 55.47 (58.57) |  |
| **Lesion location** |  |  |  |  |  |  |  |  |  |
| Frontal lobe |  |  | **0.07 (0.06)** |  | **0.01 (0.46)** |  | **0.01 (0.50)** |  | **0 (0.95)** |
| Parietal lobe |  |  | **0 (0.76)** |  | **0 (0.92)** |  | **0.03 (0.28)** |  | **0.01 (0.53)** |
| Basal Ganglia & Thalamus |  |  | **0.07 (0.07)** |  | **0.09 (0.03)\*** |  | **0.02 (0.32)** |  | **0.07 (0.06)** |
| Corpus callosum |  |  | **0.10 (0.03)\*** |  | **0.01 (0.60)** |  | **0.09 (0.03)\*** |  | **0.17 (0.004)\*** |
| Bilaterality | Unilateral | 0.33 (0.27) | **0.04 (0.20)** | 52.51 (41.01) | **0.11 (0.02)\*** | 0.42 (0.28) | **0.15 (0.006)\*** | 51.13 (55.95) | **0.11 (0.02)\*** |
|  | Bilateral | 0.23 (0.23) |  | 27.54 (25.56) |  | 0.20 (0.22) |  | 18.24 (20.85) |  |
| **Lesion extent** |  |  | **0.04 (0.16)** |  | **0.03 (0.21)** |  | **0.01 (0.64)** |  | **0.05 (0.13)** |

MM, mirror movements; CST, corticospinal tract; X, mean; SD, standard deviation.

### Combined influence of the neurological factors on mirror movements

In the **more-affected hand**, high MM-similarity was only explained by increased damage to the corpus callosum (p=0.03, R^2^=0.10, Figure 1, Table 2). MM-intensity was explained by the interaction between the CST wiring pattern and damage to the basal ganglia and thalamus (p=0.002, R^2^=0.35, Figure 2). This interaction showed that children with CSTcontra had higher MM-intensity when their basal ganglia and thalamus were more damaged. For children with CSTipsi and CSTbilat, the damage to the basal ganglia and thalamus did not influence MM-intensity in the more affected hand.

**Table 2.** Overview of the results obtained in the multiple linear regression analyses for all MM characteristics in each hand.

|  | More affected hand |  | Less affected hand |  |
| --- | --- | --- | --- | --- |
|  | MM-similarity | MM-intensity | MM-similarity | MM-intensity |
| **R<sup>2</sup>, p-value** | R<sup>2</sup>=0.10, p=0.03 | R<sup>2</sup>=0.35, p=0.002 | R<sup>2</sup>=0.59, p<0.001 | R<sup>2</sup>=0.33, p<0.001 |
| **Retained predictors** | Damage to the corpus callosum | CST wiring pattern*Damage to the basal ganglia & thalamus | Damage to the corpus callosum<br>Presence of unilateral lesions<br>CST wiring pattern*Lesion type | Damage to the corpus callosum<br>Presence of unilateral lesions |

MM, mirror movements; CST, corticospinal tract.

**Figure 1.**
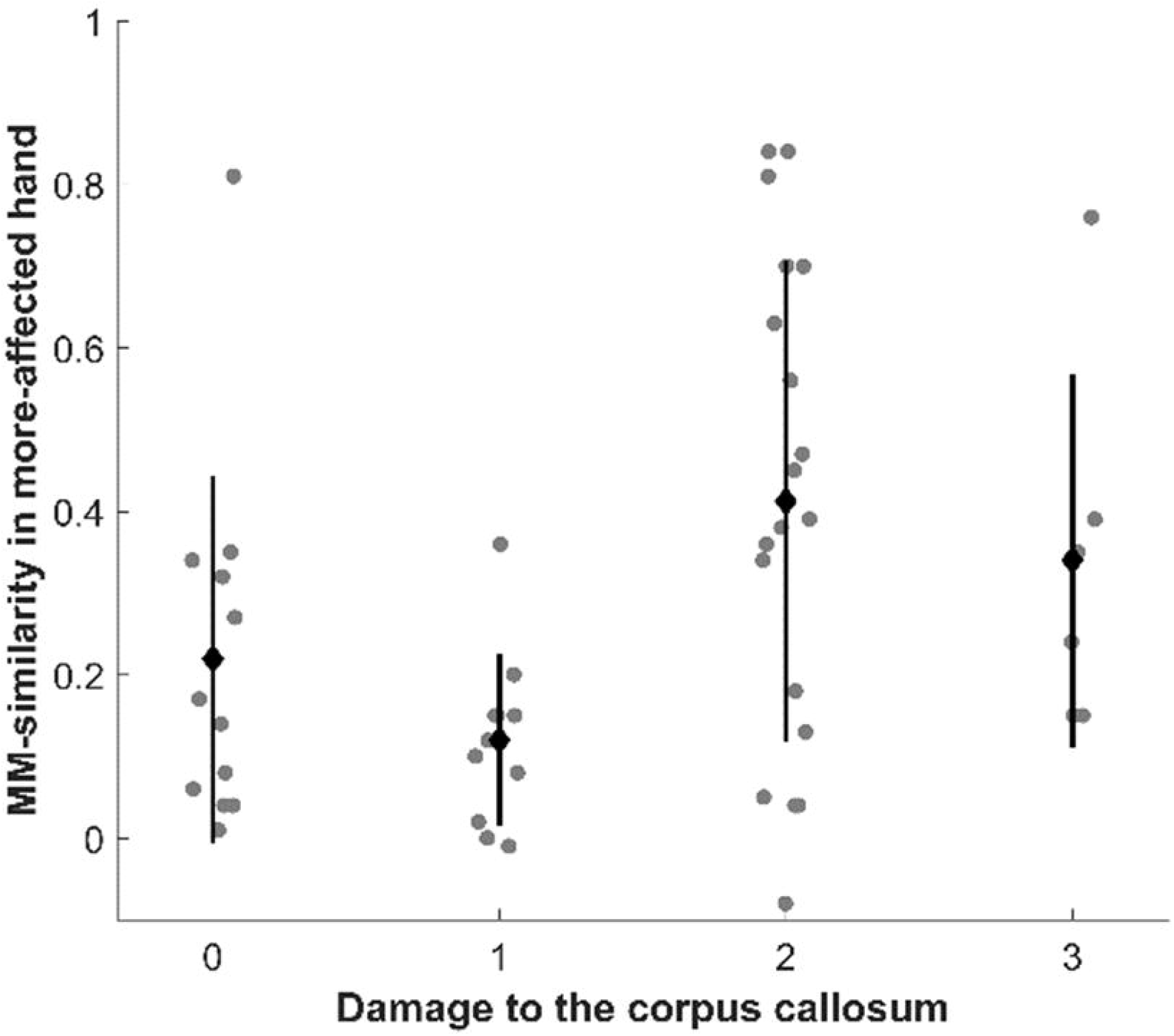
Higher damage to the corpus callosum leads to higher MM-similarity in the more-affected hand. Black dot and bar indicate mean and standard deviation, respectively.

**Figure 2.**
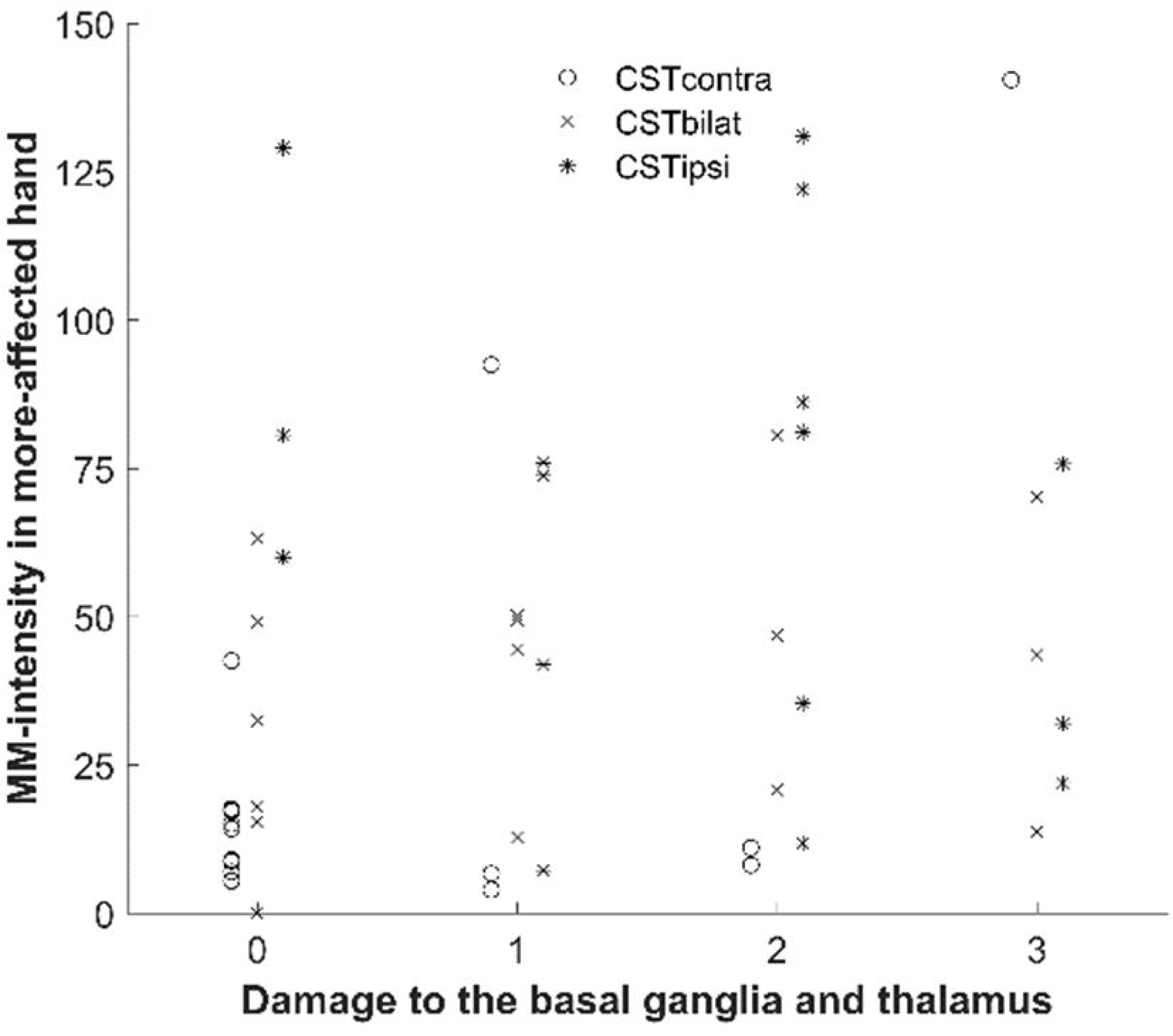
MM-intensity in the more-affected hand is explained by the interaction of the CST wiring pattern and the damage to the basal ganglia and thalamus, whereby having a CSTcontra led to higher MM-intensity when the basal ganglia and thalamus were damaged. Note that there is only one child with CSTcontra and high damage to the basal ganglia and thalamus.

In the **less-affected hand**, there were more significant influencing factors, including an interaction between the CST wiring pattern and lesion type, the damage to the corpus callosum and the presence of unilateral lesions (p<0.001, R^2^=0.59). The interaction indicated that children with a CSTcontra and CSC lesion had higher MM-similarity than those with PVL (Figure 3C). In the CSTipsi and CSTbilat groups, children with CSC lesions had lower MM-similarity compared to PV lesions. Also, children with damage to the corpus callosum and the presence of unilateral lesions showed higher MM-similarity (Figure 3A and 3B). MM-intensity was influenced by the damage to the corpus callosum and the presence of unilateral lesions (p<0.001, R^2^=0.33, Figure 4), whereby higher MM-intensity occurred with increased damage to the corpus callosum and unilateral lesions.

**Figure 3.**
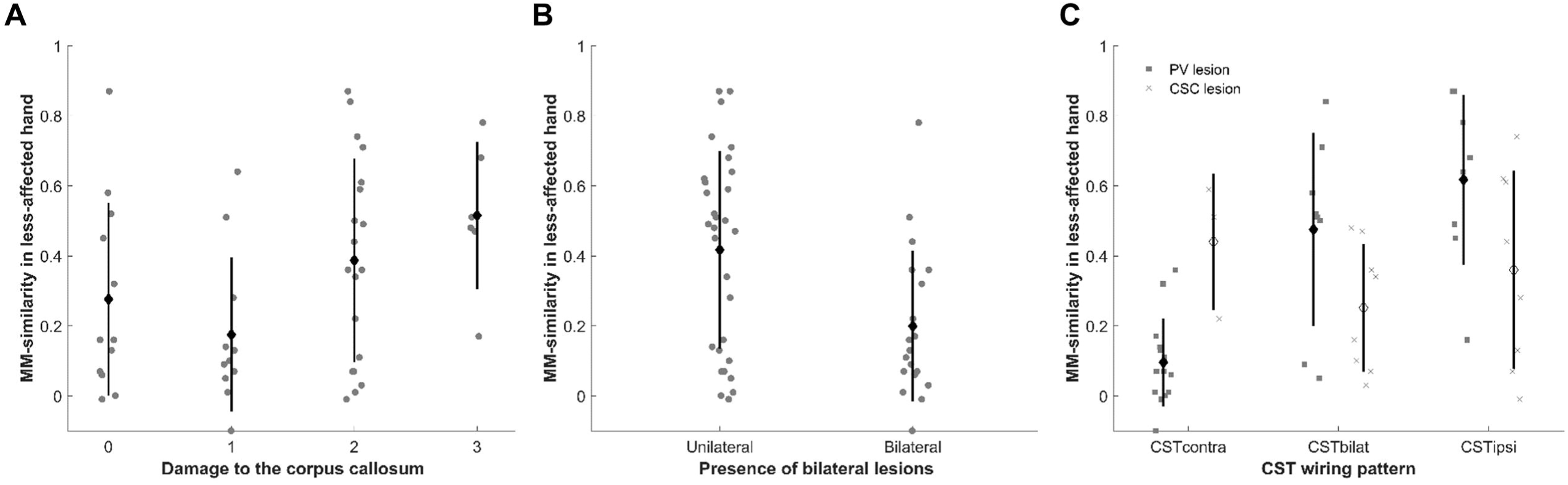
MM-similarity in the less-affected hand is explained by A) the damage to the corpus callosum (higher similarity with higher damage), B) the presence of unilateral lesions (higher similarity with unilateral lesions) and C) the interaction between lesion type and the CST wiring pattern (higher similarity in CSTcontra in CSC lesions compared to PV lesions). Black dot and bar indicate mean and standard deviation, respectively.

**Figure 4.**
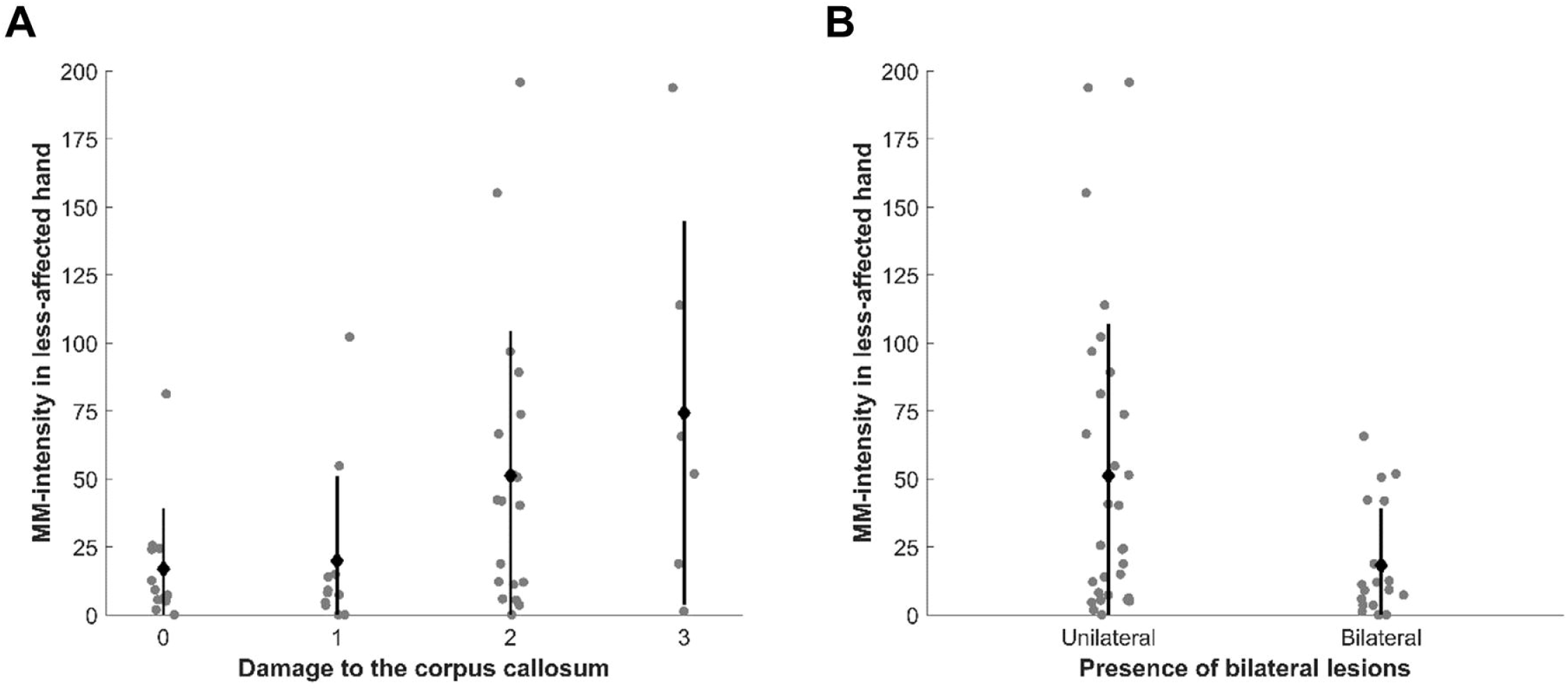
High MM-intensity in the less-affected hand was influenced by A) the increased damage to the corpus callosum and B) the presence of unilateral lesions. Black dot and bar indicate mean and standard deviation, respectively.

## Discussion

In this study, we investigated the relationship between brain lesion characteristics and CST wiring pattern on MM characteristics in each hand in children with uCP. The strength of this study lies in the use of a quantitative assessment of MM that provides different characteristics of the MM phenomenon (similarity and intensity) and the investigation of the combined value of neurological damage in a large sample size (n=49).

Previous literature, using an observational-based measure of MM evaluation (Woods and Teuber scale^5^) has demonstrated various neurological factors that influence MM occurrence in children with uCP. However, as previously highlighted, contrasting findings exist regarding the impact of the lesion type on MM occurrence^6,12^. In our study, using quantitative measurements of MM-similarity and intensity, we did not find evidence that MM characteristics differ between children with a PV or CSC lesion. Also, whilst lesion extent has been previously related to MM occurrence in children with a PV lesion^11^, our results in a larger sample size indicate that there is no relationship.

The underlying CST wiring pattern has been proposed as the main predictor of MM occurrence to such an extent that MM have been postulated to be the biomarker to evaluate the underlying CST wiring pattern without performing TMS. When investigating each factor independently, the underlying type of CST wiring was found to influence MM occurrence in our study. These simple linear regression results confirm previous study results suggesting that the presence of an ipsilateral tract (either in CSTipsi or CSTbilat wiring pattern) is related to a higher MM-similarity and MM-intensity^11–13^. However, our results also highlight the large overlap and variability between CST-wiring groups, limiting the explanatory power of the underlying CST wiring pattern. Such large variability in the CSTbilat group could be explained by a different group of neurons (i.e. different hotspot) triggering the ipsilateral and contralateral projection from the non-lesioned hemisphere, which has also been suggested to impact upper limb function^24,25^. The outcomes from this study demonstrate that there is no single factor that explains the MM phenomenon with high predictive power. Overall, our results suggest that the presence of MM cannot be used as an accurate clinical biomarker for the underlying CST wiring pattern in children with uCP.

The multifactorial interaction explaining the variability in MM-similarity in the *less-affected hand* is not so straight forward. Here, the interaction between lesion type and CST wiring pattern, damage to the corpus callosum and the presence of unilateral lesions were retained. The interaction suggested that children with CSC and CSTcontra showed higher MM-similarity in the less affected hand, whilst children with PV lesion and CSTcontra showed lower MM-similarity. Additionally, children with CSTbilat and CSTipsi who had a CSC lesion showed lower MM-similarity than those with a PV lesion. Whilst the presence of an ipsilateral projection (for both CSTbilat and CSTipsi) could explain the presence of MM, as proposed by various studies^11–13^, the high MM-similarity found in the CSTcontra group with a CSC lesion is obviously not due to the presence of an ipsilateral CST. In this case, a lack of interhemispheric inhibition could be the pathophysiological mechanism underlying MM in the less-affected hand^7^. Future studies should include neurophysiological techniques of interhemispheric inhibition to confirm this hypothesis in children with CSTcontra and CSC lesions.

Interestingly, the presence of unilateral lesions was a factor solely influencing MM in the less-affected hand. It is well known that up to 50% of the children with uCP have bilateral lesions despite clearly lateralized impairments^26^. The current results show that children with purely unilateral lesions have more MM-similarity and higher MM-intensity in the less-affected hand. These novel results suggest that the directionality of the MM (i.e. which hand mirrors and which hand drives the movement) may be driven by the presence of unilateral lesions. However, such time-related properties should be further investigated by measuring time-lag outcomes. Although this has never been shown before, it seems plausible that children with purely unilateral brain damage may have different cortico-cortical and interhemispheric connectivity compared to those with bilateral damage. The bilateral damage may trigger both hemispheres to create bypasses and additional connections to thrive beyond the damaged tissue, whilst the unilateral damage may trigger the non-lesioned hemisphere to reorganize. Supporting this hypothesis, Eyre et al (2007) showed a similar pattern of reorganization in children with uCP compared to children with bilateral lesions (potentially leading to bilateral CP)^27^.

In the *more-affected hand*, we found an interaction for MM-intensity between CST wiring pattern and damage to the basal ganglia and thalamus, indicating that children with a CSTcontra showed higher MM-intensity if their basal ganglia and thalamus were damaged. These results should be interpreted with caution and replicated in future studies as there was only one child with CSTcontra with extended damage to these subcortical structures. Previous research has shown that damage to the basal ganglia and thalamus results in poor bimanual performance (measured with the Assisting Hand Assessment)^18^, which is related to the presence of MM^6^. The basal ganglia play an important role in selecting the muscles required for a motor task, as well as for the execution of further improving already acquired motor skills^28^. In children with CSTcontra, there might be a deficiency of the basal ganglia in supporting the cortical networks to synchronously execute the movements required to perform a motor task. This hypothesis is supported by known impaired and reduced connectivity between the cortex and the basal ganglia and thalamus in children with uCP^29^. There is evidence that other brain regions besides the primary motor cortex and the corticospinal tract wiring pattern (i.e. supplementary motor network and the dorsal premotor cortex) are crucial for the performance of unilateral movements and the disruption of any part of this network will result in increased symmetrical movements^30^. Functional connectivity could shed some light in investigating the interactions between these regions. Recent studies investigating functional connectivity in children with PV lesions showed that, compared to controls, children with CST contra had increased connectivity between the primary motor cortex and the premotor cortices in the non-lesioned hemisphere^31^ whilst all children with PV lesions had decreased connectivity between the primary motor cortex and the supplementary motor area^32^. This promising technique may help in elucidating whether these brain regions influence MM in children with uCP.

Across multiple regression models, the most often retained factor was damage to the corpus callosum. The corpus callosum is a white matter bundle that acts as the main source of interhemispheric connectivity. The lack of its myelination during development is thought to be related with physiological MM in typically developing children^4^. It is plausible that the presence of a brain lesion impedes the complete myelination of the corpus callosum resulting in pathological and persistent MM due to the lack of effective communication between the hemispheres^7^. Weinstein et al (2014) showed that stronger MM were related to a lower number of transcallosal fibers^33^, suggesting that a lack of interhemispheric inhibition might be a possible mechanism underlying an increased MM-intensity in children with uCP. Future studies should focus on subparts of the corpus callosum that connect both primary motor cortices or premotor cortices^14^ or other neurophysiological techniques that can further inform causality^34^.

This study only included children with uCP resulting from PV and CSC lesions, which precludes the generalization of results to children with malformations or postnatally acquired lesions. The inclusion of multiple factors in the analyses also resulted in few children when divided into groups according to the different neurological factors. Therefore, current study results should be replicated in a larger sample. Future studies should focus on the evaluation of interhemispheric facilitation and inhibition paradigms to investigate to what extent MM occurrence depends on the interhemispheric connectivity, while taking into account the CST wiring pattern and the presence of bilateral lesions.

This is the first study to investigate MM in relation to the underlying brain lesion that used a quantitative approach to evaluate MM, allowing identification of both similarity and intensity of the MM. The evaluation of MM with the Windmill task not only gives additional information about the different MM features, but has also been shown to additionally increase the sensitivity and specificity of the MM assessment^16^. We encourage future studies to quantitatively evaluate MM providing different features of MM that can deepen our understandings on its occurrence. Current and future investigations in the underlying pathophysiology of MM will assist in the development of new treatment strategies to target the underlying mechanism.

## Conclusion

This study advances our understanding in the mechanisms of MM occurrence. Whilst the CST wiring pattern seems to be the most important factor when studied alone, it does not seem to play the largest role in influencing the occurrence of MM in uCP when other neurological factors are considered together. The underlying pathophysiology of MM in children with uCP is a complex multifactorial problem that is mainly explained by damage to the corpus callosum, basal ganglia and thalamus and is linked to unilateral lesions.

## Data Availability

Data is accessible upon request to the corresponding author

## Acknowledgements

The authors thank all the families and children who participated in this study. We also specially thank Jasmine Hoskens for her assistance during the clinical assessments.

This work is funded by the Fund Scientific Research Flanders (FWO project, grants G087213N and G0C4919N), the Special Research Fund, KU Leuven (OT/14/127, project grant 3M140230) and the Cerebral Palsy International Research Foundation (CPIRF, #R-801-11).

